# Career mentoring matters: A multi-component program for early-stage HIV investigators at the University of California, San Francisco

**DOI:** 10.64898/2026.02.24.26346718

**Authors:** Jonathan D. Fuchs, Jason S. Melo, John A. Sauceda, Joseph Watabe, Lauren Sterling, Mallory O. Johnson, Monica Gandhi

**Author notes:** Correspondence should be addressed to Jonathan D. Fuchs, MD, MPH, Center for Learning & Innovation, San Francisco Department of Public Health, 25 Van Ness Ave, Suite 500, San Francisco CA, 94102, USA.

## Abstract

**Background:** Evidence supports the key role research mentors play in bolstering the success of early stage investigators (ESI). However, there are limited data about the impact of supplemental, cross-disciplinary career mentorship and professional development opportunities for ESIs seldom included during academic training. We assessed the perceived value of this approach among post-doctoral fellows and early career faculty who participated in a multi-component career mentoring program organized by the University of California, San Francisco Center for AIDS Research (UCSF CFAR).

**Methods:** We surveyed past program participants (2005-2020), assessing demographics, current career status, perceived impact of the program, and feedback on program elements. We performed thematic analysis on open-ended responses to explore program benefits.

**Results:** Of 146 program participants contacted, 102 responded (70% response rate). Over two thirds (65%) were female, and 38% self-identified as underrepresented minority (URM) investigators. A majority of respondents now dedicate >70% of their time to research. All would recommend the program to ESI colleagues, and over 80% reported that their CFAR mentors influenced their career trajectories in several ways, including help with grant writing, linkage to researchers sparking new collaborations, and support through personal challenges or navigating conflict with primary research mentors. While 90% of URM ESIs valued advice from CFAR mentors, only a third reported receiving specific support around challenges faced as minoritized investigators.

**Conclusions:** A career mentoring program designed to complement the support offered by research mentors positively influenced the career trajectory of ESIs. Focused efforts are needed to support URM investigators who face ongoing structural barriers to success in academic settings.

## BACKGROUND

Multiple empirical studies have demonstrated the impact of formal mentoring practices on the success of post-doctoral trainees and early career faculty in academic medicine [1–3]. Skilled mentors are needed to help mentees increase both their productivity and career satisfaction [4–5]. Institutions also can play a vital role in advancing a culture of mentoring by rewarding mentoring as a formal review criterium in academic promotions. Over twenty years ago, a campus-wide workforce experience survey of faculty and staff at the University of California, San Francisco (UCSF) found that, while early-stage investigators (ESIs) praised the overall research environment, many reported inconsistencies in the mentoring experience [6]. The university responded by charging groups from across campus to put forward novel countermeasures. Our case study is an evaluation of the UCSF Center for AIDS Research (CFAR) Mentoring Program over 15 years of enrollment.

The UCSF CFAR is one of 19 domestic centers funded by the National Institutes of Health (NIH) to provide institutional support for HIV research at the intersection of basic, clinical and socio-behavioral HIV science. The CFARs harness a series of cores, including a developmental core that invests in pilot research projects proposed by ESIs. The UCSF CFAR developmental core launched a pilot mentoring program in 2005 that invited established CFAR investigators to nominate post-doctoral trainees and Assistant Professor-level faculty to join. ESIs were invited from the UCSF Schools of Medicine, Nursing, Dentistry, and Pharmacy; from UCSF graduate programs; and from the CFAR’s affiliated research institutions (e.g., the Gladstone Institutes, the San Francisco Department of Public Health) to participate in a novel two-year pilot program. At its heart, the program matched each ESI mentee with a volunteer mid-career or senior faculty member who did not serve as a primary research mentor. In fact, program leadership sought to match ESIs with faculty with complementary and non-competing research interests. Designated as CFAR “career mentors”, these faculty were asked to review ESIs’ individual development plans and offer relevant insights on traditional mentoring topics (e.g., manuscript and grant writing) as well as non-traditional ones (e.g., life/work balance, negotiating faculty positions). Content and frequency of meetings were driven by mentee demand. If mutually beneficial, mentees and career mentors could elect to extend their relationships beyond the two years. The program also included monthly professional development workshops and an annual ESI symposium to showcase their work for UCSF’s extended HIV research community.

Grounded in social cognitive career theory [7], the CFAR posited the program would impact both the quality of mentoring and foster a mentoring culture by investing in complementary, unbiased support for ESIs that would accelerate their progress and feelings of inclusion and belonging. Evaluation of the pilot program found mentees highly valued the professional networking from the program [8]. Early findings also catalyzed several program enhancements including the introduction of specific aims “lightning rounds”, which are peer review sessions in advance of twice-yearly pilot award and HIV/AIDS designated NIH grant deadlines. A full day, interactive ESI leadership retreat was added, and to increase their visibility, the program launched an annual research excellence award ceremony for ESIs who were nominated by the CFAR community.

As one of the longest standing, formalized career mentoring programs for ESIs conducting multi-disciplinary HIV research, the UCSF CFAR mentoring program sought to gather additional input from past program participants on the expanded program model and explore how the program influenced their career trajectories. Based on these findings, we share key lessons with stakeholders designing and implementing ESI-focused mentoring programs.

## METHODS

### Program Description

Components of the UCSF CFAR mentoring program are outlined in **Figure 1**. Annually, investigators from across the CFAR enterprise nominated all HIV-focused post-doctoral fellows and recently-appointed assistant professors to participate in the CFAR Mentoring Program. During the pilot phase, Program Directors assigned mentees to senior faculty who were not involved in their HIV research. Subsequently, as the program evolved, mentees submitted preferences for these career mentors with whom they would meet on a quarterly or more frequent basis to share challenges and opportunities. Whether assigned or chosen by the mentee, the CFAR career mentor was asked to adhere to a set of expectations, including reviewing the mentee’s individual development plan and meeting with the mentee regularly.

**Figure 1:**
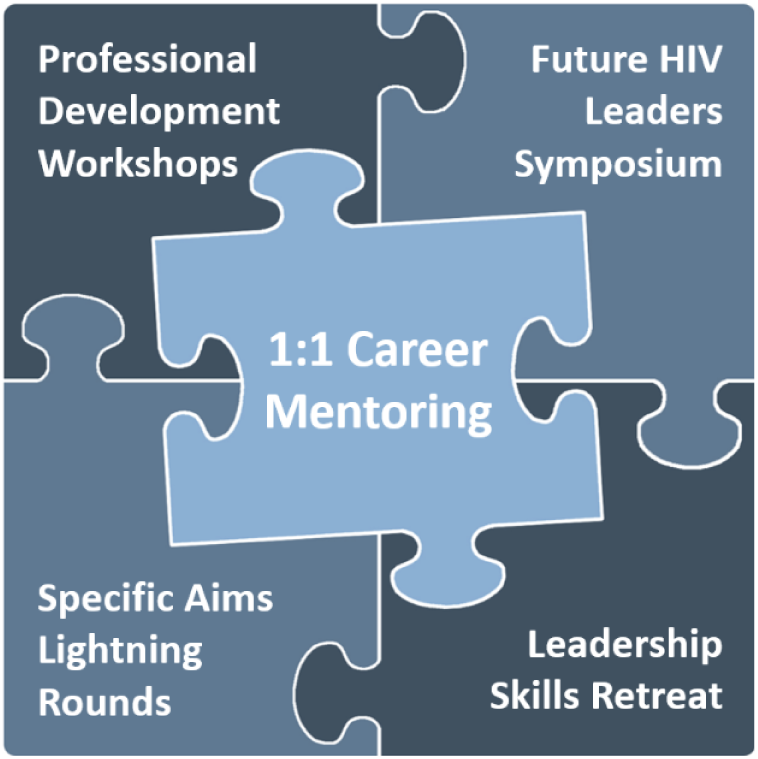
Core components of the UCSF CFAR Career Mentoring Program.

The program also held monthly professional development workshops covering topics prioritized by the mentees including how to build and sustain a team, how to negotiate at various stages of training, time management, manuscript writing, grant writing, how to build a budget, the mentor-mentee relationship structure, unconscious bias, how to give and receive feedback, and research resources available across the CFAR enterprise. Three mentees nominated by the CFAR community are selected yearly to receive excellence awards in basic, clinical and social-behavioral sciences and to present their research at a designated CFAR seminar. Other ESIs are asked to present at an annual Future Leaders in HIV Research Symposium to increase ESI visibility. After the two-year program pilot, input from ESIs inspired new programming to build grant writing capacity through specific aims lightning rounds peer review sessions and the addition of an interactive ESI leadership retreat covering a range of topics such as increasing emotional intelligence, adopting different leadership styles, mentoring others, and effectively managing conflict. As mentees became mentors themselves, they were offered the opportunity to participate in a two-day CFAR Mentoring of Mentors Workshop, described elsewhere. [9–12].

### Participants and procedures

All individuals who participated as mentees in the CFAR Mentoring Program between 2005 and 2020 were eligible for this evaluation. From February through April of 2021, a total of 146 unique program participants were invited by email to complete a ten-minute web-based survey (Supplemental File 1) developed for this study via Qualtrics (Provo, UT). Weekly reminders were sent to encourage survey completion. In addition, participants received a US$10 gift code for an online retailer and were entered into a raffle for an additional one of two randomly awarded US$100 gift codes to incentivize survey completion.

### Survey content

The survey’s primary aim was to determine the long-term impact of program participation on mentees and their career trajectory. Additionally, the survey gathered data on all program elements and how the program, as a whole, succeeded and could be improved to support future cohorts of ESIs.

The survey included questions about demographics, professional disciplines, current positions, and if they have mentored others through their career. Status as an underrepresented minority (URM) trainee was defined as being a member of an NIH-described historically underrepresented racial and ethnic minority group [13], and/or a gender-identity minority (i.e., transgender or gender non-binary-identified individuals).

Participants were asked to score their satisfaction with core aspects of the CFAR Mentoring Program (1-Very Satisfied to 4-Very Dissatisfied Likert-type scale), the value they attributed to professional development workshops (1-Very Helpful to 4-Not Helpful At All Likert-type scale), and the perceived long-term impact of the program on career success (1-Strongly Agree to 4-Strongly Disagree Likert-type scale). In all three scoring domains, participants were given additional response options in the case that the program aspect they were scoring did not apply to them (e.g., Did Not Participate, Do Not Remember). All 4-point Likert scales were dichotomized for analysis.

These items were supplemented with an open-ended question that asked participants to describe what the most valuable aspect was of the mentoring relationship they had with their CFAR career mentor. We performed a thematic analysis [14] of these narrative responses, in which team members (JSM and JDF) reviewed answers, discussed and finalized themes, and selected illustrative quotes to showcase each theme.

### Ethics statement

This assessment was conducted as part of program evaluation for the UCSF CFAR, and thus ethics approval was not required.

## RESULTS

### Respondent characteristics

Of 146 program participants contacted, 102 (70%) responded to the survey. The characteristics of these participants are shown in **Table 1**. Approximately two thirds were female, 25% Asian, Native Hawaiian, or Pacific Islander, 8% Black/African American, and 6% reported Latino/a/x ethnicity. In total, 39 (38%) participants self-identified as a racial/ethnic or sexual and/or gender identity URM investigator. At the time of survey completion, median age was 39.5 years with a range from 30 to 66 years. More than one in five reported being the first to attend college in their immediate family. Respondents came from a variety of research disciplines—social/behavioral science (25%), clinical science (24%), basic science (20%), translational science (16%), epidemiology (15%), and medical education (2%). Respondents were largely post-doctoral fellows (68%) whereas 20% were assistant professors during their time in the program.

**Table 1.** Trainee demographics and background during program participation (N = 102)

|  |  | **N** | **%** |
| --- | --- | --- | --- |
| **Gender Identity<sup>1</sup> (n=95)** | Female | 62 | 65.3 |
|  | Male | 31 | 32.6 |
|  | Gender minority | 2 | 2.1 |
| **Race<sup>1,2</sup> (n=95)** |  |  |  |
|  | Black/African American | 7 | 7.7 |
|  | Asian | 20 | 22.0 |
|  | Native Hawaiian or Pacific Islander | 3 | 3.3 |
|  | White | 60 | 65.9 |
|  | Other | 5 | 5.5 |
| **Latino/a/x or Hispanic Ethnicity<sup>1</sup> (n=94)** | Yes | 6 | 6.4 |
|  | No | 88 | 93.6 |
| **Any underrepresented minority (URM) characteristic identified** | Yes | 39 | 38.2 |
|  | No | 63 | 61.8 |
| **First in family to attend college<sup>1</sup> (n=94)** | Yes | 21 | 22.3 |
|  | No | 73 | 77.7 |
| **Research discipline in the program** | Basic Science | 20 | 19.6 |
|  | Clinical Science | 24 | 23.5 |
|  | Translational Science | 16 | 15.7 |
|  | Social/Behavioral Science | 25 | 24.5 |
|  | Epidemiology | 15 | 14.7 |
|  | Medical Education | 2 | 2.0 |
| **Training stage during the program** | Post-doctoral Fellow | 69 | 67.6 |
|  | Assistant Professor | 20 | 19.6 |
|  | Associate Professor | 1 | 1.0 |
|  | Staff Scientist | 4 | 3.9 |
|  | Other | 8 | 7.8 |
|  | **Median (IQR)** | **Range** |  |
| **Current age<sup>1</sup> (n=84)** | 39.5 (36.0 – 44.0) | 30 – 66 |  |

<sup>1</sup>Respondents with no answer (i.e., skipped, preferred not to answer) are not included in the total for these questions.
<sup>2</sup>For race, Ns do not add up to total, as individuals could select more than one option.

### Career trajectories

Trainee career trajectory and outcome measures are shown in **Table 2**. Nearly four in five respondents currently work in an academic setting, and the majority dedicate more than 70% of their work to research. Given our interest in cultivating mentorship through this program, we also assessed their experience as mentors. We found respondents currently mentor an average of 4 mentees; at the time of the survey, only 11% are not mentoring.

**Table 2.** Program outcomes and reported career trajectories (N = 98)

|  | N | % |
| --- | --- | --- |
| **Career advice from their CFAR mentor influenced career trajectory:** |  |  |
| Among all trainees | 79 | 80.6 |
| Among underrepresented minority (URM)<sup>1</sup> trainees (N=39) | 36 | 92.3 |
| **Among those that responded yes, their career was influenced in the following ways:** |  |  |
| Helped plan for a K award | 32 | 40.5 |
| Helped with applications for CFAR or NIH funding | 31 | 39.2 |
| Linked to researchers that sparked new collaborations | 30 | 38.0 |
| Offered support when dealing with a personal challenge | 17 | 21.5 |
| Helped get a paper accepted for publication | 15 | 19.0 |
| Encouraged pursuit of a desired job in industry, government, non-profit, etc. | 14 | 17.7 |
| Helped address conflict with a primary mentor | 12 | 15.2 |
| Helped get an abstract accepted for presentation | 10 | 12.7 |
| Helped with applications for other funding | 10 | 12.7 |
| Changed the focus of research | 9 | 11.4 |
| **Among URM trainees that responded yes, their career was also influenced in the following ways:** |  |  |
| Received support as a URM<sup>1</sup> investigator | 10 | 27.8 |
| **Current Employer** |  |  |
| Academic | 77 | 78.6 |
| Government | 6 | 6.1 |
| Industry | 6 | 6.1 |
| Non-profit | 7 | 7.1 |
| Other | 2 | 2.0 |
| **Percent of current job dedicated to research** |  |  |
| >70% | 59 | 60.2 |
| 50-70% | 9 | 9.2 |
| 30-50% | 8 | 8.2 |
| 10-30% | 12 | 12.2 |
| <10% | 10 | 10.2 |
|  | **Mean(SD)** | **Range** |
| **Number of people to whom the respondent currently mentors<sup>2</sup>:** | 4 (5) | 0 – 24 |

<sup>1</sup>Underrepresented minority trainees are defined as respondents who self-reported as identifying with a racial, ethnic, or gender minority.
<sup>2</sup>Mentees may include faculty, post-doctoral fellows, graduate students, medical students, undergraduates or others. 11% are not mentoring currently.

Most mentees reported that the career advice they received from their mentor during the program positively influenced their career trajectory (81%), and this was even more strongly reported among URM mentees (92%). Among all mentees who responded favorably to this question about CFAR mentors, the most common ways mentors influenced their careers was through help with planning for a K award (41%), helping with applications for CFAR or NIH funding (39%), and establishing connections to researchers that sparked new collaborations (38%). Among URM trainees who responded yes that CFAR mentors positively influenced their career, fewer than a third of these respondents (28%) responded affirmatively to the statement that “my CFAR mentor offered support to me specifically as a URM investigator”.

### Program feedback

Respondent feedback on program components and outcomes was overwhelmingly positive, as shown in **Figure 2**. Trainees were especially satisfied with program leadership and monthly workshops overall (≥99% satisfied), followed by meeting fellow ESIs, mentor assignment, and completing individual development plans (≥90% satisfied). All stated they would recommend the program to any post-doctoral fellows or early career faculty considering participation. Respondents also largely agreed that they benefitted from the program (97%), that the program advanced a culture of mentoring (94%), and that the program helped them achieve career goals (87%). Most respondents also agreed the program made them both better mentors and mentees (86% and 85%, respectively).

**Figure 2.**
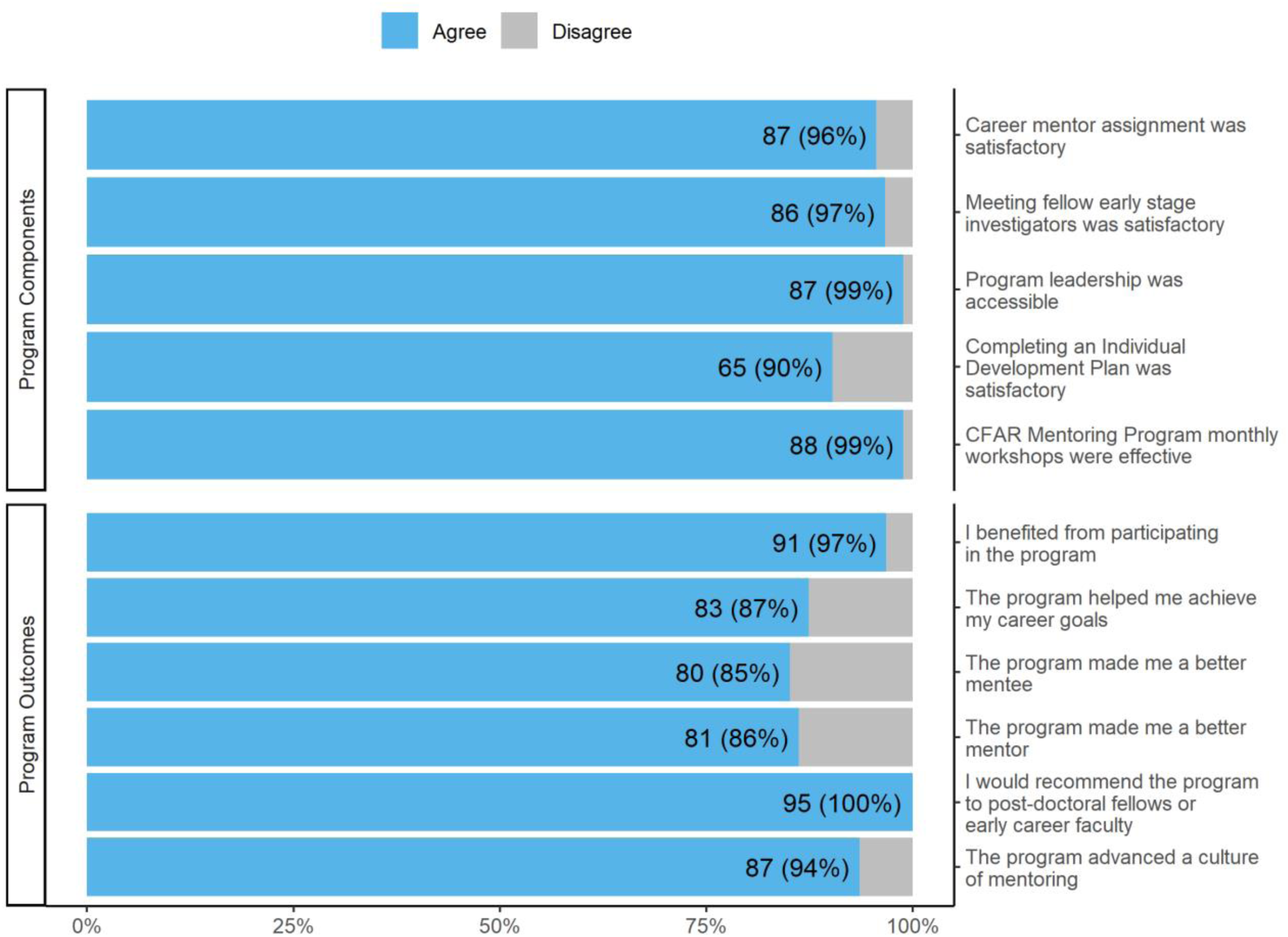
Trainee reflections on program components and outcomes (N = 95)^1^. ^1^Respondents with no answer (i.e., skipped, preferred not to answer) are not included in the total for these questions. Therefore, the total for each question varies.

Respondents also provided feedback on specific workshops, as shown in **Figure 3**. Among those who attended each workshop and provided feedback, ≥90% of attendees found the workshops helpful 17 out of 19 times. Five workshops were reported helpful by 100% of respondents (e.g., Coaching and Giving Feedback, Grant Writing, Leadership and Teamwork, Specific Aims Lightning Rounds, and Unconscious Bias), whereas a lower proportion rated the overview of research happening across the multiple institutions that comprise the CFAR highly.

**Figure 3.**
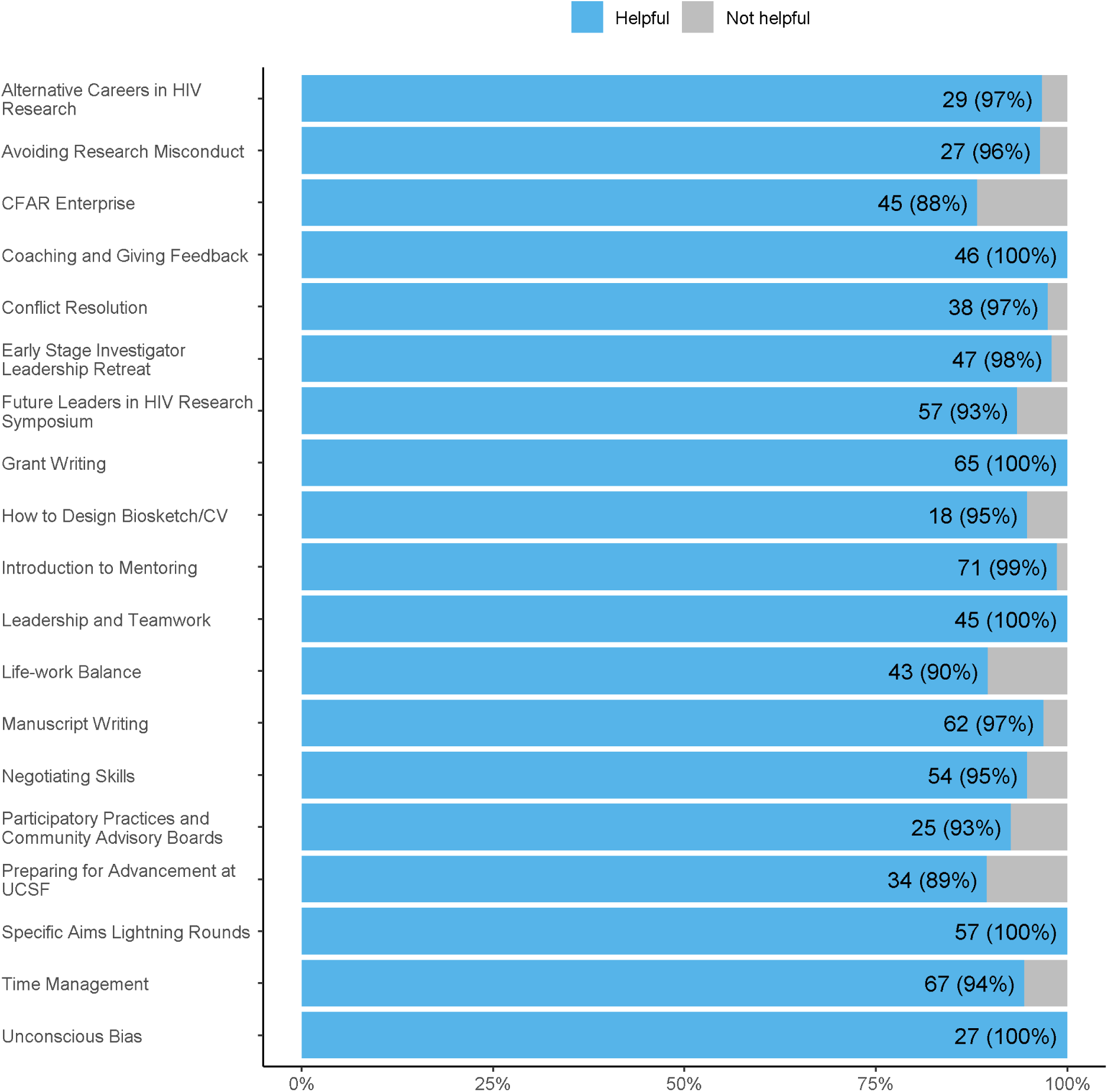
Trainee reflections on specific program workshops (N = 94)^1^. ^1^Respondents with no answer (i.e., skipped, did not attend, does not remember) are not included in the total for these questions. Therefore, the total for each workshop evaluation varies.

### Qualitative comments

Thematic analysis of narrative responses allowed us to better understand what trainees valued most about their relationship with their mentor during the CFAR Mentoring Program and how these relationships impacted them both during the program and over the long-term. Several key themes emerged. Illustrative quotes from respondents with their corresponding demographic characteristics, discipline, and trainee or faculty status at the time of the program are offered below.

The first and most prominent theme was the professional guidance and career navigation offered by senior mentors. Former trainees highlighted their mentors’ assistance navigating academic research, determining a career path, and succeeding in one’s chosen professional setting.

> *“I learned a great deal about what is valued in academic research settings as productive, which helped me evaluate whether this setting is conducive to a healthy work-life balance, my mental health, and whether the pace and incentives of research actually help address the health issues we research.”*
>
> — —Female, Black/African American, Social/Behavioral Science, Post-Doctoral Fellow

> *“The relationship is an enduring one. A primary mentoring relationship that was foundational to my early career success and continues to be highly influential in my ongoing career trajectory.”*
>
> — —Gender Minority, White, Social/Behavioral Science, Assistant Professor

The next most common theme - personal and confidential support - was derived from responses highlighting the importance of garnering emotional support from an outside career mentor without having to bring such concerns to a direct supervisor.

> *“Having another faculty member who understood the trials I was going through as a trainee and in my lab. Knowing someone else was cheering me on was extremely helpful, particularly when I faced hardships.”*
>
> — —Female, White, Basic Science, Post-Doctoral Fellow

> *“The ability to talk frankly about challenges balancing an unreasonable workload and ideas to problem solve that.”*
>
> — —Male, White, Social/Behavioral Science, Assistant Professor

A substantial number of respondents commented on the value of mentor feedback on grant applications and other scientific writing projects which comprised the third key theme.

> *“He helped support me when I had some challenges in transitioning to writing a K grant.”*
>
> — —Female, Asian, Translational Science, Associate Professor

> *“His incredible expertise and support provided valuable clinical research input which helped me get the UCSF RAP [intramural pilot research] award.”*
>
> — —Female, White, Basic Science, Post-Doctoral Fellow

Networking opportunities emerged as its own theme, as many respondents pointed to building new connections via their CFAR mentor’s professional network.

> *“[My mentor] has done an outstanding job of connecting me with other researchers, sparking new directions for my research.”*
>
> — —Female, White, Social/Behavioral Science, Post-Doctoral Fellow

> *“Continuing to build my academic and professional network in the Bay Area.”*
>
> — —Female, Asian, Epidemiology, Post-doctoral Fellow

The fifth and final theme focused on the benefits of mentorship outside their own research discipline. Trainees often found having a mentor with a different background or not directly involved in their research helped them gain new perspectives on their work.

> *“For me as a basic scientist the outside perspective coming from a clinical scientist helped me to gain a big picture view of my work.”*
>
> — —Male, White, Basic Science, Post-Doctoral Fellow

> *“Many of my mentors are specific to a research project. It has been hugely beneficial to have a career mentor who sees the forest for the trees and can advise me on appropriate strategies and directions moving forward.”*
>
> — —Male, Black/African American, Epidemiology, Post-Doctoral Fellow

## DISCUSSION

The UCSF Center for AIDS Research mentoring program was launched in 2005 and has demonstrated great promise in providing complementary cross-disciplinary career mentoring for post-doctoral and early career faculty in HIV research. We conducted our first longitudinal assessment of over 100 previous program participants to explore its perceived value during and several years after they were matched to senior career mentors and participated in a wide range of professional development opportunities, including monthly workshops, grant reviews, symposia and leadership training. Our evaluation showed that most former mentees in the program continue to engage in HIV research in university and non-university settings, and were often serving as mentors themselves. The vast majority of respondents reported that being part of the program favorably influenced their career trajectories and all would recommend it to trainees during this critical, formative stage of their careers.

Mentorship from senior CFAR faculty was perceived as the most valuable component of the program, and for many, had a clear influence on their research paths by providing career advice and input on grants and other scientific writing. Consistent with the program pilot evaluation, our assessment showed that participants highly valued the active role career mentors played in helping to expand their professional networks [8]. Independent career mentors offered key emotional support to many respondents, and in some cases, helped them manage conflict with their primary research mentor(s). Several respondents also reported that their CFAR mentors encouraged them to explore career opportunities outside of academia such as government and industry which are key alternative settings to conduct research given the limited number of university faculty positions and an increasingly constrained funding environment.

Through this assessment, we were committed to understanding the perspectives of our URM-identified mentees who comprised 38% of the sample. This group found the career advice they received from the program helpful, at a rate even higher than the cohort overall. However, among these respondents, fewer than a third shared they received advice specifically focused on their minoritized identities. The ability to mentor across identity differences is an established mentoring core competency [15] and continues to be a high priority as published reports expose the ongoing threat to retaining URM investigators, particularly those from historically underrepresented racial and ethnic minoritized backgrounds in academia [16–17]. Structural factors associated with differential retention include, but are not limited to, lower award rates to Black/African American investigators pursuing NIH R01 funding [18], experiences of interpersonal racism in the workplace [19], and a well-described “minority tax” placed on URM investigators who are frequently tapped to lead or support institutional diversity, equity, and inclusion efforts and who are highly sought after by URM trainees for mentoring. In many cases, URM faculty engage in these activities, often without sufficient protected time for these pursuits [20].

Addressing these barriers require structural solutions. Since 2012, our CFAR has hosted an annual two-day, in-person Mentoring the Mentors training which has spotlighted these systemic issues for national cohorts of participating mid-career and senior mentors in HIV research. This workshop helps address how mentors can effectively mentor across differences, address implicit biases, and advocate effectively for their individual mentees and for institutional change [9–12]. In addition, we launched a new program to provide designated support, mentoring and advocacy for racial and ethnic URM ESIs in HIV research at UCSF. This program (led by JS), offers additional scaffolding for URM ESIs including expanded direct advocacy, networking, professional development opportunities, retention support, and other assistance as requested by mentees (e.g., intervening to help resolve conflicts). In January of 2022, our CFAR launched an inter-CFAR URM Working Group which created a URM investigator-led vision for HIV research and aims to translate this vision across the national network of CFARs.

There are limitations of this evaluation. First, we recognize that this assessment cannot establish a causal relationship between program participation and longitudinal career outcomes without a suitable comparison group. This was methodologically infeasible as virtually all early-stage HIV researchers affiliated with the CFAR are offered the opportunity to participate and choose to do so, and random assignment to an experimental condition would be both impractical and poorly accepted [8]. We did, however, query former program participants directly about the perceived impact and through categorical and open-ended survey responses, found that most strongly valued the career mentorship received and shared how it supported their research aspirations—an explicit goal of the mentoring program. Second, while we observed a high response rate to this survey, it is possible that non-respondents decided not to take it because they pursued career paths other than research and felt a longitudinal assessment was less applicable to them. Another possible reason was that non-respondents may not perceive the same value of the program as survey respondents. We found, however, that 10% of individuals who did respond to the survey were not engaged in any research activities in their current role but were still motivated to share their insights. In addition, since the program’s inception, our annual participant evaluation completed by nearly all mentees has consistently shown high satisfaction ratings with program components making it less likely that non-respondents were less enthusiastic about the program overall, reducing the chance of bias. Finally, results may not be generalizable to academic settings outside the U.S. where career trajectories for research and mentoring expectations may differ.

## CONCLUSIONS

In summary, a multi-component strategy that offered career mentoring and a structured professional development curriculum for early-stage HIV investigators seems to positively influence their career trajectories and contribute to their success in advancing HIV research. Program participants highlighted that the CFAR Mentoring program answered the university’s original charge 20 years ago to strengthen the mentoring culture which, at that time, was inconsistent. To support the long-term success of ESIs, academic research programs should consider enlisting the support of well-trained mentors who are committed to fostering diversity so they can participate in cross-disciplinary career mentoring and augment primary research mentoring relationships. As the UCSF CFAR has now started a designated URM ESI mentoring program, ongoing evaluation of this program is underway. Future work is needed to assess the experiences of our senior career mentors and identify ways we can further support them as they invest in the next generation of HIV research leaders.

## DECLARATIONS

### Ethics approval and consent to participate

This study is based on program evaluation materials and did not involve human subjects as defined by the federal regulations summarized in 45 CFR 46.102(e). The Institutional Review Board at the University of California San Francisco therefore determined that it did not require IRB oversight.

### Consent for publication

This study did not include human subjects as defined by the federal regulations summarized in 45 CFR 46.102(e); it did not include identifiable information for any individual.

### Availability of data and materials

Due to confidentiality within a relatively small group, data sharing is limited, but available upon request.

### Competing interests

The authors declare no competing interests. The content is solely the responsibility of the authors and does not necessarily represent the official views of the National Institutes of Health or the San Francisco Department of Public Health.

### Funding

This work was supported by grants P30AI027763 (MG), K24MH087220 (MOJ), and R25DA043441 (JDF and JAS).

### Authors’ contributions

JDF led preparation of the article. JDF designed the study. JSM led the quantitative data analysis and development of tables and figures. JDF and JSM co-led the qualitative, thematic analysis. JDF, JSM, JAS, JW, LS, MOJ, and MG all contributed significantly to close review and revision of the article.

## Acknowledgements

We would like to thank our UCSF CFAR Mentoring program participants for their reflections on this program, and our CFAR faculty who have ably served as career mentors since 2005.

## List of Abbreviations

HIV: Human immunodeficiency virus
UCSF: University of California, San Francisco
CFAR: Center for AIDS Research
ESI: Early Stage Investigator
Clinical Trial Number: Not applicable.

## Notes

### Competing Interest Statement

The authors have declared no competing interest.

### Funding Statement

This work was supported by grants P30AI027763, K24MH087220, and R25DA043441.

