## Supplemental File 1 for "Career mentoring matters: A multi-component program for early-stage HIV investigators at the University of California, San Francisco"

### Default Question Block

Dear ${m://FirstName},

During our 17th year of the CFAR Mentoring Program, we'd like to take stock in how participation in this program affected you and your career trajectory. Looking back, we'd appreciate your thoughts on what worked particularly well and what could be improved as we aim to support future cohorts of early career investigators.

This survey will take **approximately 10 minutes** to complete, and we will send you a $10 Amazon gift code upon completion. We will also enter you for a drawing for one of two $100 Amazon gift codes (on top of your guaranteed $10 code). Your responses are confidential and all data will be reported in aggregate.

Please click on the arrow at the bottom of the page to begin.

You participated in the program from ${e://Field/Start Year} to ${e://Field/End Year}.
**Dr. ${e://Field/Mentor}** was your CFAR career mentor during the program. For the those of you who participated in the early years of the program and had two CFAR mentors, please describe your experience with the CFAR career mentor with whom you had the most contact.

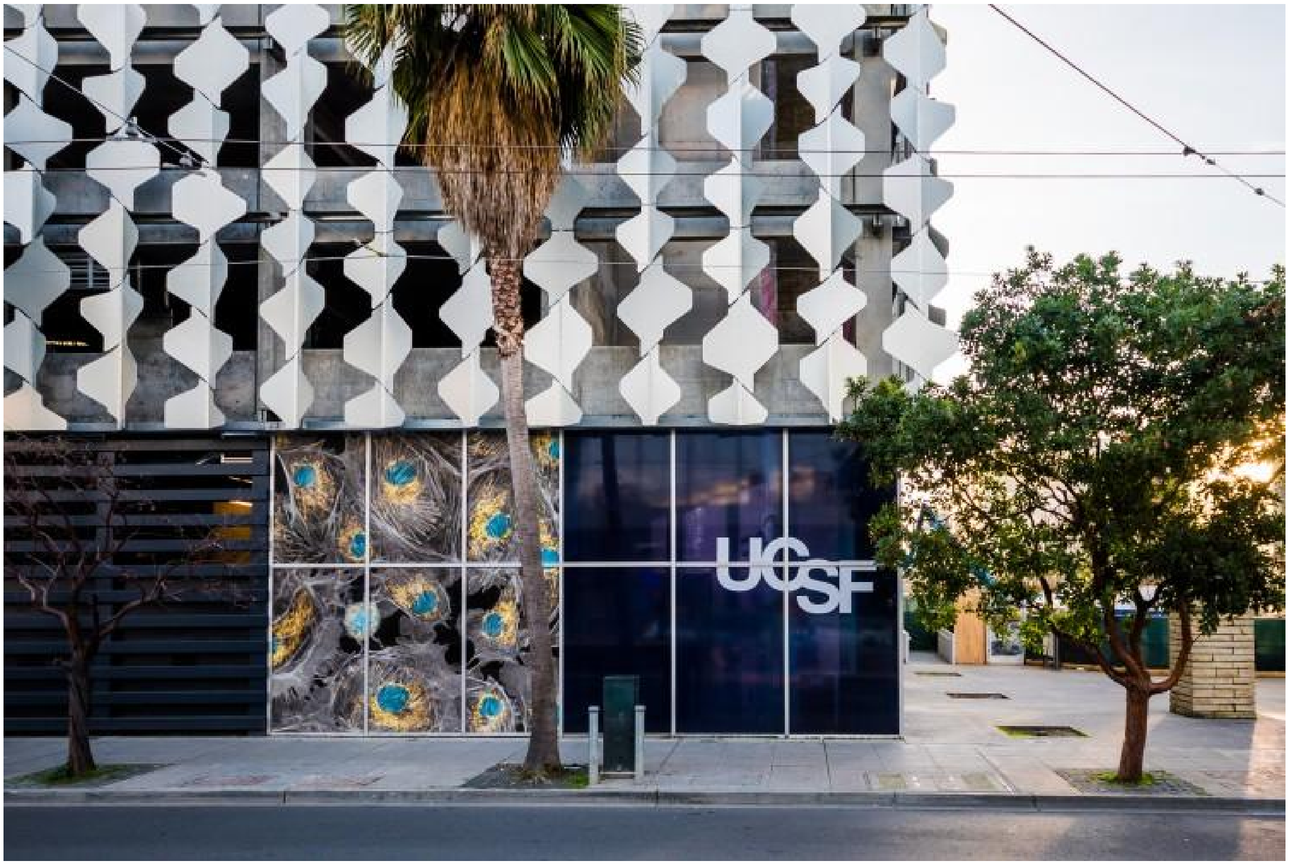


CFAR Center for AIDS Research
University of California San Francisco
Gladstone Institute of Virology & Immunology

### Part 1: Your background when you were a CFAR mentoring program mentee

What was your stage of training when you participated?

Post-doctoral fellow

Assistant Professor

Associate Professor

Staff scientist

ID fellow

TAPS fellow

[ ] Other (please describe)

What was your research discipline during your time in the program?

Basic science

Clinical science

Translational science

Social/behavioral

Epidemiologic

Policy

Medical education

Other (please describe)

During the program, approximately how many times did you meet with your CFAR career mentor?

0

1-2

3-4

5-6

More than 6

Did you continue to meet with your CFAR career mentor after your first year in the program?

Yes

No

CFAR Center for AIDS Research
University of California San Francisco
Gladstone Institute of Virology & Immunology

### Part 2: Where are you now?

Where are you currently employed?

- [ ] Academic institution (please describe)
- [ ] Industry (please describe)
- [ ] Government (e.g. public health)
- [ ] Non-profit (please describe)
- [ ] Other (please describe)

What is the zip code of your primary institution?

What percentage of your current job is dedicated to research?

<10%

10-30%

30-50%

50-70%

>70%

How many people do you mentor currently?

Faculty

Post doctoral fellows

Graduate students

Medical students

Undergraduates

Others

I am not mentoring at this time

Since you participated in the program (and including those years), how many peer reviewed papers have you published and grants have you received? Please summarize below, or -preferably- upload your cv or biosketch (below).

0 Peer reviewed papers did you publish as the primary author or coauthor?

0 NIH-funded grants did you procure as the Principal or Co-principal investigator?

0 Foundation Grants or other grants (federal or non-federal)? What was the total $ amount?

Please upload CV or biosketch here:

CFAR Center for AIDS Research
University of California San Francisco
Gladstone Institute of Virology & Immunology

### Part 3: Reflections on your CFAR mentoring experience

What do you remember most about your participation in the CFAR mentoring program?

Did the career advice you received from your CFAR mentor influence your career trajectory?

Very much

Somewhat

Not at all

If very much or somewhat, how so? (check all that apply)

Helped me get an abstract accepted for presentation

Helped me get a paper accepted for publication

Helped me plan for a K award

Helped me with applications for CFAR or NIH funding (e.g., RAP pilot award, other NIH award)

Helped me with applications for other funding (e.g., foundation funding)

Linked me to researchers that sparked new collaborations

Changed the focus of my research

Encouraged me to follow my desire to leave academia and pursue a job in industry, government, non-profit or other setting

Helped me address a conflict I was having with a primary research mentor

Supported me as I was dealing with a personal challenge

Offered support to me as an underrepresented minority (URM) investigator

Other (please describe)

Is there advice you received you didn’t follow, but wished you had?

Yes, please describe

No

Don't Know

What was the most valuable aspect of the mentoring relationship you had with your CFAR career mentor?

CFAR Center for AIDS Research
University of California San Francisco
Gladstone Institute of Virology & Immunology

### Part 4: Reflections on Program Components

How satisfied were you with these core aspects of the CFAR Mentoring program?

| | Very satisfied | Satisfied | Not satisfied | Very dissatisfied | Didn't participate |
|---|---|---|---|---|---|
| Being assigned a career mentor | O | O | O | O | O |
| Getting to know fellow early stage investigators | O | O | O | O | O |
| Access to program leadership | O | O | O | O | O |
| Completing an Individual Development Plan (IDP) | O | O | O | O | O |

| | Very satisfied | Satisfied | Not satisfied | Very dissatisfied | Didn't participate |
|---|---|---|---|---|---|
| CFAR Mentoring Program monthly workshops | O | O | O | O | O |

Which professional development workshops were valuable in supporting your personal/professional growth?

| | Very helpful | Helpful | Not helpful | Not helpful at all | Didn't participate | Do not remember |
|---|---|---|---|---|---|---|
| Introduction to mentoring | O | O | O | O | O | O |
| CFAR Enterprise | O | O | O | O | O | O |
| Time management | O | O | O | O | O | O |
| Manuscript writing | O | O | O | O | O | O |
| Grant writing | O | O | O | O | O | O |
| Alternative Careers in HIV Research: Industry and Non-Profit | O | O | O | O | O | O |
| Coaching and Giving Feedback – Helpful Tips | O | O | O | O | O | O |
| Leadership and Team Work | O | O | O | O | O | O |
| Negotiating Skills - Getting what you want | O | O | O | O | O | O |
| Conflict resolution | O | O | O | O | O | O |
| Avoiding research misconduct | O | O | O | O | O | O |
| How to design biosketch/CV | O | O | O | O | O | O |
| Preparing for advancement at UCSF | O | O | O | O | O | O |
| Life-work balance | O | O | O | O | O | O |
| Good participatory practices and working with community advisory boards | O | O | O | O | O | O |

| | Very helpful | Helpful | Not helpful | Not helpful at all | Didn't participate | Do not remember |
|---|---|---|---|---|---|---|
| Future Leaders in HIV Research Symposium, including annual research excellence awards | O | O | O | O | O | O |
| Specific Aims Lightning Rounds | O | O | O | O | O | O |
| Early stage Investigator Leadership Retreat (usually held in June) including several topics such as the Myers Briggs personality inventory and leadership styles | O | O | O | O | O | O |
| Unconscious Bias | O | O | O | O | O | O |

Looking back, is there is a workshop topic that wasn’t covered, but would have been helpful to you at that stage in your career?

Yes, please describe

No, I felt the curriculum was complete

How much do you agree with the following statements?

| | Strongly Agree | Agree | Disagree | Strongly disagree |
|---|---|---|---|---|

| | Strongly Agree | Agree | Disagree | Strongly disagree |
|---|---|---|---|---|
| I benefited from my participation in the CFAR Mentoring Program | ○ | ○ | ○ | ○ |
| My participation in the CFAR Mentoring Program helped me achieve my career goals | ○ | ○ | ○ | ○ |
| Participating in the program helped me be a better mentee | ○ | ○ | ○ | ○ |
| Participating in the program helped me be a better mentor | ○ | ○ | ○ | ○ |
| I would recommend the CFAR Mentoring Program to post-doctoral fellows or early career faculty | ○ | ○ | ○ | ○ |
| The CFAR mentoring program advanced a culture of mentoring in the HIV community at UCSF and affiliated organizations | ○ | ○ | ○ | ○ |

Any advice for program organizers on how the CFAR Program can be improved to support early stage investigators?

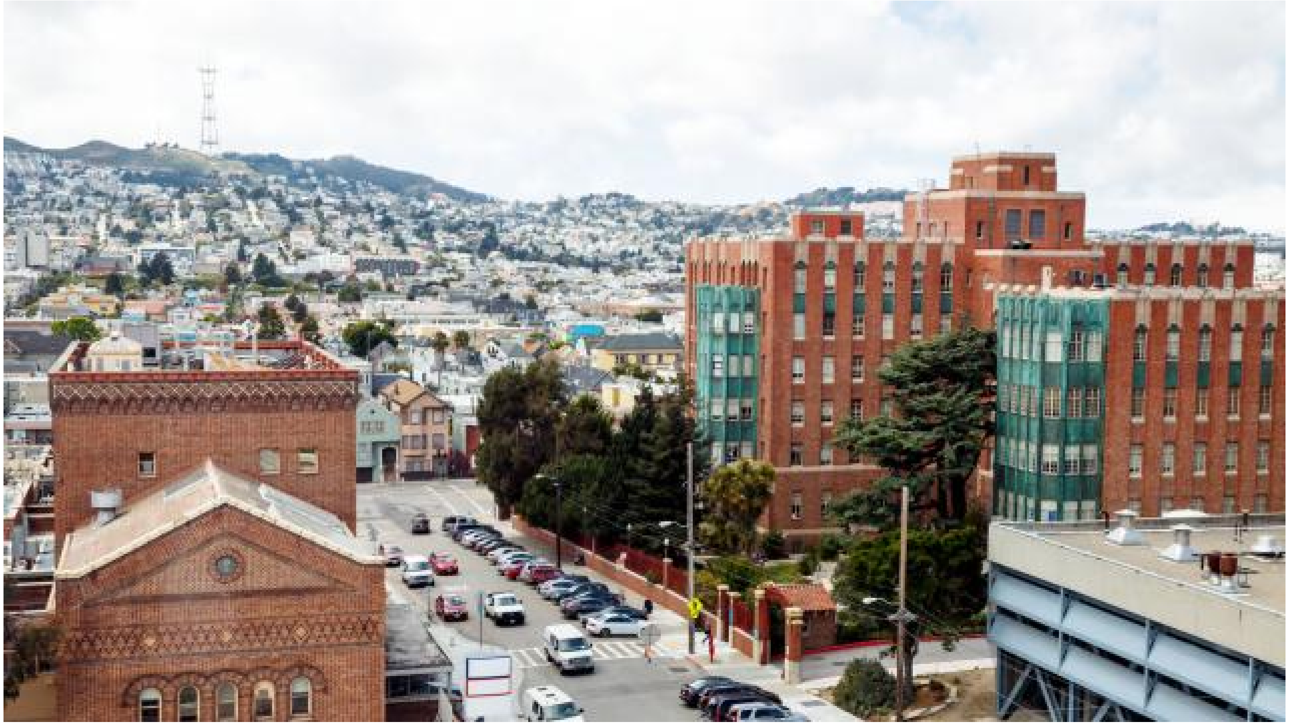

CFAR Center for AIDS Research
University of California San Francisco
Gladstone Institute of Virology & Immunology

### Part 5: About you

What is your year of birth?

[ ] Year

Prefer not to answer

Do you identify as (select all that apply)

Male

Female

Transgender, non-binary, or other gender minority identity

Other

Prefer not to answer

Do you identify as Latino/a/x or Hispanic?

Yes

No

Prefer not to answer

Do you identify as? (Check all that apply)

Black or African-American

American Indian/Alaskan Native

East Asian

South Asian

Native Hawaiian or other Pacific Islander

White

Other (please specify)

Prefer not to answer

Are you the first person from your immediate family to attend college?

Yes

No

Prefer not to answer

Testimonials from participants of our CFAR Mentoring Program are the best form of advertisement for the program. Are there comments about your experience with the CFAR Mentoring Program that we might use to support future funding and advertisements for ESIs considering participation? We welcome your quotes here:

If we use this quote, would you be okay with our listing your name and current affiliation? (e.g, Dr. Jane Smith, University of The World School of Medicine says “your quote here”). There would be listed for noncommercial purposes such as on our website, NIH progress reports and funding applications, presentations, etc.

Yes, please feel free to include my name (with the following considerations)

No, please do not use my name

Not applicable

Powered by Qualtrics
